# Sex-specific risk of smoking for abdominal aortic aneurysm and exploration of potential mechanism: meta-analysis and prospective cohort study

**DOI:** 10.1101/2024.11.08.24316961

**Authors:** Paul Welsh, Anna-Louise Pouncey, Janet T Powell

## Abstract

**Background:** Smoking is the strongest modifiable risk factor for abdominal aortic aneurysm (AAA). This study aims to confirm whether smoking is a stronger risk factor in women than men and identify contributory reasons, including inflammation, for any sex-specific difference observed.

**Methods:** Systematic review and meta-analysis, conducted according to PRISMA guidance (Prospero registration (CRD2024586609)). Data sources were Medline, Embase, and CENTRAL. Population-based studies reporting risk of AAA, adjusted for age and cardiovascular risk factors, for women versus men, were included. These were complemented by data from the UK Biobank (UKB) cohort, which also were assessed for sex-specific effects of smoking on incident atherosclerotic cardiovascular disease (ASCVD). Results: Meta-analysis of 6 studies (including UKB), 2001-2024) showed that the relative risk ratio of current versus never-smokers for incident AAA in women versus men was 1.78 [95%CI 1.32, 2.38]. Comparison of the sex-specific relative risks of current smoking and number of cigarettes/day were similar in the UKB cohort and these risks were much higher for AAA than for ASCVD, but the risks of pack-years were similar. Sex-specific risks of current smoking for AAA were not significantly modified by inflammatory markers (including C-reactive protein, alkaline phosphatase and white blood cell count), lung function or physical activity. Stopping smoking reduced the risk of AAA by almost half in both sexes. Conclusions: The risk of developing AAA by current smokers is almost twice as high in women versus men. Inflammation was not a major modifier and other reasons for the disparity must be sought.

## Introduction

Smoking is a stronger risk factor for abdominal aortic aneurysm (AAA) than for most other common forms of cardiovascular disease(1). Three studies have previously indicated the effect of smoking on AAA development appears to be stronger in women than men (2-5). With the knowledge that AAA is up to 6 times more common in men(5, 6), and that smoking amongst adult men are only slightly higher than in women(7), such observations may seem surprising. However, it is recognized that men and women may smoke differently and for different reasons. Men and women choose different types of cigarettes(8), and appear to inhale cigarettes differently(9). Neuroimaging studies support the activation of differential brain pathways, with men smoking primarily for the reinforcing or reward effects of nicotine, whereas women tend to smoke to regulate mood or in response to cigarette-related cues(2).

The culmination of cultural and physiological differences may result in different health outcomes. There is strong evidence that the risks of smoking (10) for the development of other diseases are stronger in women, particularly the risk of chronic obstructive pulmonary disease (COPD)(11, 12). A modest effect for atherosclerotic cardiovascular disease (ASCVD) also has been demonstrated but no underlying mechanisms have been identified. The very strength of the association of smoking with AAA may offer a better possibility of identifying mechanisms. Therefore, the aim of this study is to consolidate the evidence for sex-specific differences in AAA through systematic review and meta-analysis and explore UK Biobank data for underlying mechanisms. Since there are sex-specific effects of smoking on ASCVD and chronic lung disease, the hypothesis that inflammation might be the underlying reason was explored.

## Methods

### Systematic review

The systematic review was registered in PROSPERO (CRD2024586609) and was conducted and is reported using Preferred Reporting Items for Systematic reviews and Meta-Analyses (PRISMA) statement (https://www.prisma-statement.org/). (See Supplemental Figure S1.) Using ProQuest Dialog™, a search strategy for MEDLINE, EMBASE and Cochrane Central Register of Controlled Trials (CENTRAL) using a combination of controlled vocabulary (MeSH terms and free text terms,) was formulated with the final search conducted on 31 August 2024. The MeSH terms used were smoking OR cigarettes OR tobacco AND sex AND abdominal aortic aneurysm, with free text terms of pack-years, population, screening and longitudinal studies. There were no date restrictions. Only publications which included an English language abstract were included and the reference lists of any reviews or systematics reviews identified were searched manually. If necessary, study authors were contacted for further information. The essential inclusion criterion was that the risk of AAA was compared for current smokers and never smokers for men and women separately. Other inclusion criteria were population-based studies, risk results presented with 95% confidence intervals and either adjusted at least for age or providing data to enable age-adjustment. Eligible studies could provide adjusted odds ratios (ORs), hazard ratios (HRs) or Relative risks (RRs) and their 95% confidence interval (CI) or provide data to enable estimation of such a risk. Risk of bias was assessed using the ROBINS-E tool (https://www.riskofbias.info/welcome/robins-e-tool; Supplemental Figure S2). ORs and HRs, (assuming constant hazards over time,) were converted to RRs using the baseline risk or the probability of the event in the control group and the delta method for approximation of the variance. Exclusion criteria included reports only in languages other than English, reports of hospital studies or AAA repairs and case-control studies with <200 AAA cases. Data were extracted by 2 independent reviewers onto Excel spreadsheets. Meta-analysis of relative risk for men and women, and of relative risk ratios for women-to-men, were conducted using the inverse variance method with the DerSimonian-Laird estimator for tau^2^, the Jackson method for the confidence interval of tau^2^, and Hartung-Knapp adjustments for random effects, with graphical representation using forest plots. A p value of <0.05 was considered significant. The Forest plots also show the prediction interval, which is the range in which the point estimates of 95% of future studies are expected to fall (assuming that the effect sizes are normally distributed). Heterogeneity was assessed using a chi^2^ test and the I^2^ statistic. Further “leave one out” and “influence analyses, using Baujat plots, externally standardized residuals, DFFITS value, Cook”s distance, covariance ratio, leave one out tau^2^ and Cochran”s Q values to identify studies to exclude, were used to examine the robustness of findings. Analyses were conducted in R Studio (http://www.rstudio.com) using the DMetar package(13).

### UK Biobank data and cohort selection

The original data used in this study are available via UK Biobank (https://www.ukbiobank.ac.uk/), subject to necessary approvals. UK Biobank is a large population-based cohort study of 502 188 participants ranging in age from 37 to 73 years, recruited between 2006 and 2010(14, 15). All participants underwent an assessment at 1 of 22 centres across England, Scotland, and Wales, where touch-screen questionnaires recorded health and lifestyle information, participants underwent an interview for health conditions and medication use, and a wide range of biological measurements were taken. Covariates in the analysis included age, sex, ethnicity (white, black, South Asian, other), systolic and diastolic blood pressures, (both the average of two measurements,) Townsend deprivation score (a postcode based measure of socioeconomic deprivation), body mass index (BMI), weight, height, body fat percentage from bioimpedance, diabetes (type 1 or type 2), International Physical Activity Questionnaire (IPAQ)-measured physical activity (low, moderate or high), chronic kidney disease, baseline atherosclerotic cardiovascular disease, and lung function (FEV1 (forced expiratory volume in 1 second) and FVC (forced vital capacity)). Medications included blood pressure lowering medications, cholesterol lowering medications, and antiplatelet medications obtained for self-report or nurse interview). Blood biomarkers were obtained according to a standardised protocol.

The primary exposure of interest was smoking status, categorized as never, ex-smoker, and current smoker. Smoking habit was also explored using number of cigarettes smoked per day (zero in non-smokers, imputing unknown smokers as the median per day, and ten cigarettes a day amongst pipe and cigar smokers) and using pack-years of smoking (UK Biobank data field 20161). These were analysed to assess whether the sex interaction held after accounting for smoking intensity and cumulative exposure.

Participants were excluded if they self-reported aortic aneurysm, aortic aneurysm rupture, or aortic dissection (UK Biobank data field 20002) or first occurrence of aortic aneurysm or aortic dissection was recorded as occurring before recruitment (UK Biobank data field 131382) (total n excluded =591. We conducted a complete case analysis in 382762 participants with complete data for the primary adjustment model.

UK Biobank received ethical approval from the North West Multi-Centre Research Ethics Committee (REC reference: 11/NW/03820). All participants gave written informed consent before enrolment, in accordance with the principles of the Declaration of Helsinki. This project was performed under UK Biobank project approval No. 71392.

#### Outcomes

All clinical data in the hospital inpatient data were coded according to the World Health Organization”s (*International Classification of Diseases, Tenth Revision* [*ICD-10*]) codes. All operations and procedures were coded according to the Office of Population, Censuses and Surveys: Classification of Interventions and Procedures codes (OPCS-4). Dates and causes of death were obtained from death certificates held by the NHS Information Centre for participants from England and Wales and the NHS Central Register Scotland for participants from Scotland.

Implementation of the United Kingdom National Health Service (NHS) screening policy for AAA in men aged 65 or over was complete in most parts of the United Kingdom by late 2009.^17^ Participants with complete data for primary adjustment variables (*vide infra*) were followed up until 31st August 2022 in Scotland, 31st October 2022 in England, and 31st May 2022 in Wales.. The outcome of interest was defined as previously using first hospital inpatient diagnosis of AAA or death from AAA (both based on ICD-10 codes I71.3 or I71.4), or an AAA-related surgical procedure. A secondary outcome was ASCVD defined as death from coronary heart disease or stroke (I20-I25, I60-I64), or hospitalisation for myocardial infarction or stroke (I21-I22, I60-I64).

### Statistical methods for UK Biobank data

Rate of AAA was obtained from crude estimates per 10,000 person years stratified by smoking status and sex. Descriptive statistics were generated for baseline characteristics, stratified by AAA and ASCVD outcomes, using means, standard deviations and t-tests for normally distributed continuous variables, medians and inter-quartile intervals and rank sum tests for skewed variables, and counts, percentages chi-square tests for categorical variables.

Cox proportional hazards regression models were used to evaluate the association between smoking status, sex, and outcomes, with AAA and ASCVD. Interaction terms between sex and smoking-related variables were included in the models to assess potential effect modification. Models were adjusted for the following confounders: age, ethnicity, Townsend deprivation score, diabetes status, total cholesterol, HDL cholesterol, blood pressure medication use, statin use, baseline ASCVD, and chronic kidney disease. Subsequently, there were two sequential sensitivity analyses adjusting for other potential variables which might modify the sex-specific effect identified. These included systolic blood pressure, diastolic blood pressure, BMI, BMI interaction with diabetes status, body fat percentage, HbA1c, markers of inflammation (alkaline phosphatase (ALP), alanine aminotransferase (ALT), aspartate aminotransferase (AST), gamma-glutamyltransferase (GGT), CRP and white cell count) and factors associated with thrombosis (platelet count and antiplatelet medication use). Second, an extended model additionally included neutrophil-to-lymphocyte ratio (NLR), lung function (FEV1 and FVC) and physical activity (IPAQ), with sex interactions for FEV1 and FVC. This latter extended model had a lower sample size due to missing data on lung function and physical activity. Analyses by number of cigarettes smoked per day and pack-year history were further investigated smoking habits. Stratified analyses by CRP levels were performed to further assess the role of inflammation. All analyses were for complete cases and conducted using Stata version 18 (StataCorp, College Station, TX).

### Data availability

Data used for the systematic review can be shared. Additional results from UK Biobank data, including all the Cox model data can be shared but source data must be obtained directly by application to UK Biobank.

## Results

### Systematic review and meta-analysis

Systematic review of the literature, to 31st August 2024, identified 421 potential studies, which after screening of the title and abstract left 31 potential studies. This reduced to 5 reports after review of the full report and data extraction(3-5, 16, 17). In addition, a search was conducted for UK Biobank studies for potentially available information. One paper was identified although it did not contain sex-specific analyses(18). The PRISMA flow diagram and risk of bias assessment are shown in Supplemental Figures S1 and S2 and details of these studies are provided in Table 1, with a total of 1050810 men and 1771349 women. Overall, studies had a low-medium risk of bias.

**Table 1.** Studies included in the meta-analysis or risk of abdominal aortic aneurytm in current smokers versus never smokers.

| Author<sup>ref</sup> | Year Country | Men N= | Risk of AAA | 95%CI | Women N= | Risk of AAA | 95%CI | Adjusted for age & | Risk of Bias |
| --- | --- | --- | --- | --- | --- | --- | --- | --- | --- |
| Carter<sup>4</sup> | 2024 USA & UK | 818616 | RR 7.3 | 6.4,8.2 | 1513943 | RR 15.0 | 13.2,17.0 | BMI, BP, lipids | medium |
| Jahangir<sup>3</sup> | 2015 USA | 7727 | HR 3.4 | 2.0,5.9 | 11815 | HR 9.2 | 5.0,17.0 | Race, BMI, BP, socioeconomic factors | medium |
| Koncar | 2024 Serbia | 2972 | OR 2.88 | 1.90,4.36 | 974 | OR 4.17 | 1.46,11.96 | BMI, HT, alcohol use | medium |
| Singh | 2001 Norway | 2962 | OR 7.4 | 3.7,14.7 | 2434 | OR 5.8 | 2.9,11.6 | BMI, lipids, HT, fibrinogen | low |
| Stackelberg<sup>2</sup> | 2024 Sweden | 42596 | HR 6.5 | 5.4,8.0 | 35550 | HR 11.0 | 7.4,16.3 | Education, waist circumference, diabetes, HT, hypercholesterolemia | low |
| UK Biobank | 2024 UK | 176837 | HR 3.78 | 3.38,4.17 | 206633 | HR 6.60 | 5.36,8.13 | Confounders listed above | low |

Abbreviations used: BMI body mass index, BP blood pressure, HR hazard ratio, HT hypertension, OR odds ratio, RR relative risk

For men, the pooled relative risk of AAA in current smokers versus never smokers was 4.83 [95%CI 3.0,7.62], with high heterogeneity (I^2^=94%). For women, the pooled relative risk of AAA of current smokers versus never smokers was 8.44 [95%CI 5.25,13.92], also with high heterogeneity (I^2^=90%). The pooled relative risk ratio, women to men, is shown in Figure 1. The relative risk ratio was 1.78 [95%CI 1.32,2.38], again with high heterogeneity (I^2^=96.2%). However, all findings were robust on “leave one out” in influence analyses. (See Supplemental Figures S3, S4.) Overall, these data consistently indicate that the relative risk for development of AAA associated with current smoking is almost twice as high in women as men.

**Figure 1.**
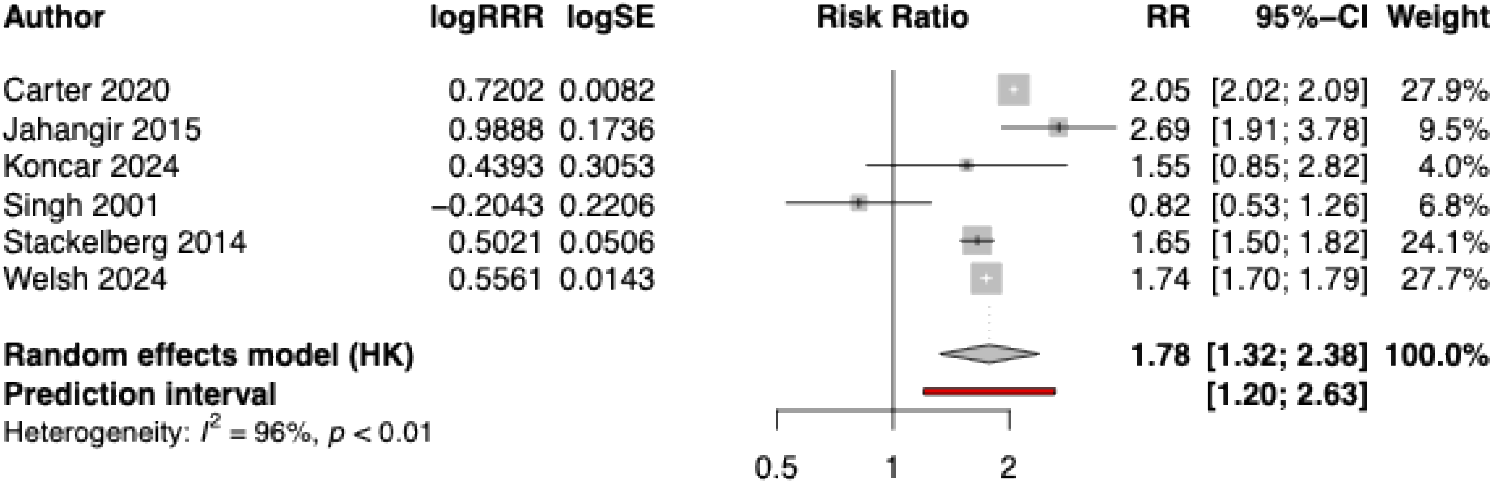
Forest plot of relative risk ratios (women to men) of risk of Abdominal aortic aneurysm in current versus never smokers

### Comparison of the sex-specific risks of smoking associated with incident AAA and ASCVD in the UK Biobank cohort

The population included 176837 men and 206633 women, with incident AAA diagnosed in 1793 and 393 respectively, with baseline characteristics shown in Table 2. Women had much lower pack-year histories of smoking than men. Other cardiovascular risk factors also differed by sex, with higher use of prevention medications in men than women. Men also had higher blood pressures, whilst women had higher serum cholesterol and CRP concentrations. Over a median 13.4 years follow-up (IQI 12.9-14.3) the rate of AAA per 10,000 person-years was 7.7 (95%CI 7.4-8.1) in men and 1.4 (95%CI 1.3-1.6) in women.

**Table 2.**
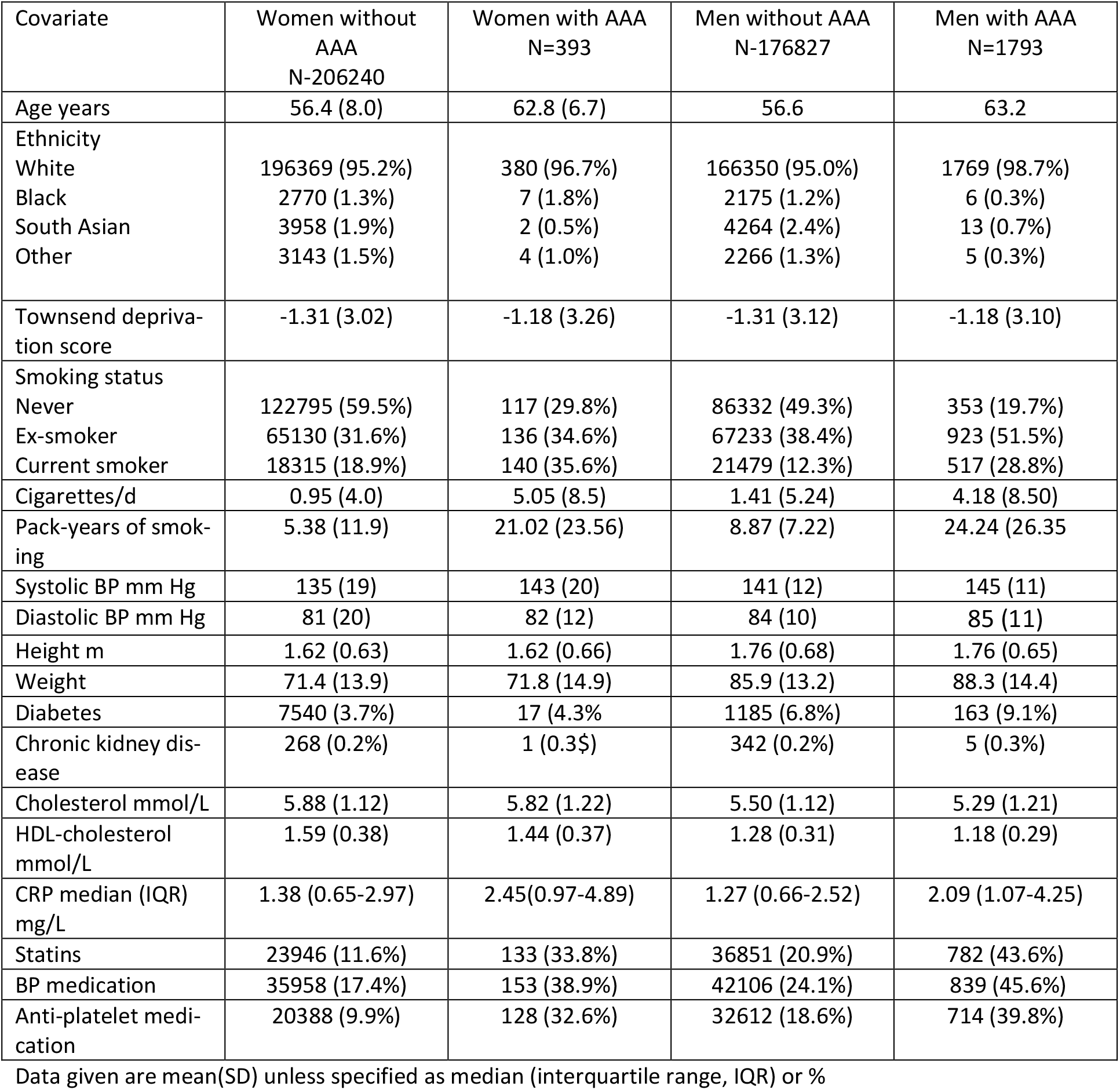
Baseline characteristics of women and men reporting incident abdominal aortic aneurysms during follow up in the UK Biobank Cohort.

| Covariate | Women without AAA<br>N=206240 | Women with AAA<br>N=393 | Men without AAA<br>N=176827 | Men with AAA<br>N=1793 |
| --- | --- | --- | --- | --- |
| Age years | 56.4 (8.0) | 62.8 (6.7) | 56.6 | 63.2 |
| Ethnicity |  |  |  |  |
| White | 196369 (95.2%) | 380 (96.7%) | 166350 (95.0%) | 1769 (98.7%) |
| Black | 2770 (1.3%) | 7 (1.8%) | 2175 (1.2%) | 6 (0.3%) |
| South Asian | 3958 (1.9%) | 2 (0.5%) | 4264 (2.4%) | 13 (0.7%) |
| Other | 3143 (1.5%) | 4 (1.0%) | 2266 (1.3%) | 5 (0.3%) |
| Townsend deprivation score | -1.31 (3.02) | -1.18 (3.26) | -1.31 (3.12) | -1.18 (3.10) |
| Smoking status |  |  |  |  |
| Never | 122795 (59.5%) | 117 (29.8%) | 86332 (49.3%) | 353 (19.7%) |
| Ex-smoker | 65130 (31.6%) | 136 (34.6%) | 67233 (38.4%) | 923 (51.5%) |
| Current smoker | 18315 (18.9%) | 140 (35.6%) | 21479 (12.3%) | 517 (28.8%) |
| Cigarettes/d | 0.95 (4.0) | 5.05 (8.5) | 1.41 (5.24) | 4.18 (8.50) |
| Pack-years of smoking | 5.38 (11.9) | 21.02 (23.56) | 8.87 (7.22) | 24.24 (26.35) |
| Systolic BP mm Hg | 135 (19) | 143 (20) | 141 (12) | 145 (11) |
| Diastolic BP mm Hg | 81 (20) | 82 (12) | 84 (10) | 85 (11) |
| Height m | 1.62 (0.63) | 1.62 (0.66) | 1.76 (0.68) | 1.76 (0.65) |
| Weight | 71.4 (13.9) | 71.8 (14.9) | 85.9 (13.2) | 88.3 (14.4) |
| Diabetes | 7540 (3.7%) | 17 (4.3%) | 1185 (6.8%) | 163 (9.1%) |
| Chronic kidney disease | 268 (0.2%) | 1 (0.3%) | 342 (0.2%) | 5 (0.3%) |
| Cholesterol mmol/L | 5.88 (1.12) | 5.82 (1.22) | 5.50 (1.12) | 5.29 (1.21) |
| HDL-cholesterol mmol/L | 1.59 (0.38) | 1.44 (0.37) | 1.28 (0.31) | 1.18 (0.29) |
| CRP median (IQR) mg/L | 1.38 (0.65-2.97) | 2.45(0.97-4.89) | 1.27 (0.66-2.52) | 2.09 (1.07-4.25) |
| Statins | 23946 (11.6%) | 133 (33.8%) | 36851 (20.9%) | 782 (43.6%) |
| BP medication | 35958 (17.4%) | 153 (38.9%) | 42106 (24.1%) | 839 (45.6%) |
| Anti-platelet medication | 20388 (9.9%) | 128 (32.6%) | 32612 (18.6%) | 714 (39.8%) |

Data given are mean(SD) unless specified as median (interquartile range, IQR) or %

**Table 3.** The risk of current versus never smoking on incident AAA stratified by CRP concentration. Results are given for the primary adjusted analyses.

| Sex | All<br>n=392952 | CRP<2 mg/L<br>n=247637 | CRP ≥2 mg/L<br>n=135125 |
| --- | --- | --- | --- |
|  | HR [95% CI] | HR [95% CI] | HR [95% CI] |
| women | 6.60 [5.36,8.13] | 5.11 [3.61,7.23] | 7.49 [5.73,0.81] |
| men | 3.78 [3.38,4.17] | 3.07 [2.54,3.59] | 3.83 [3.31,4.36] |
| Sex interaction | 0.57 [0.45,0.72] | 0.60 [0.41,0.88] | 0.51 [0.38,0.69] |

The unadjusted rates for AAA incidence per 10,000 person-years in men and women in never smokers, ex-smokers and current smokers are shown in Figure 2; rates of AAA were higher in smokers in both sexes, but the absolute risk of AAA was always higher in men regardless of smoking. The relative harms of current smoking appeared higher in women, whereas the benefits of stopping smoking appeared similar (almost 50% reduction in rate) in men and women.

**Figure 2.**
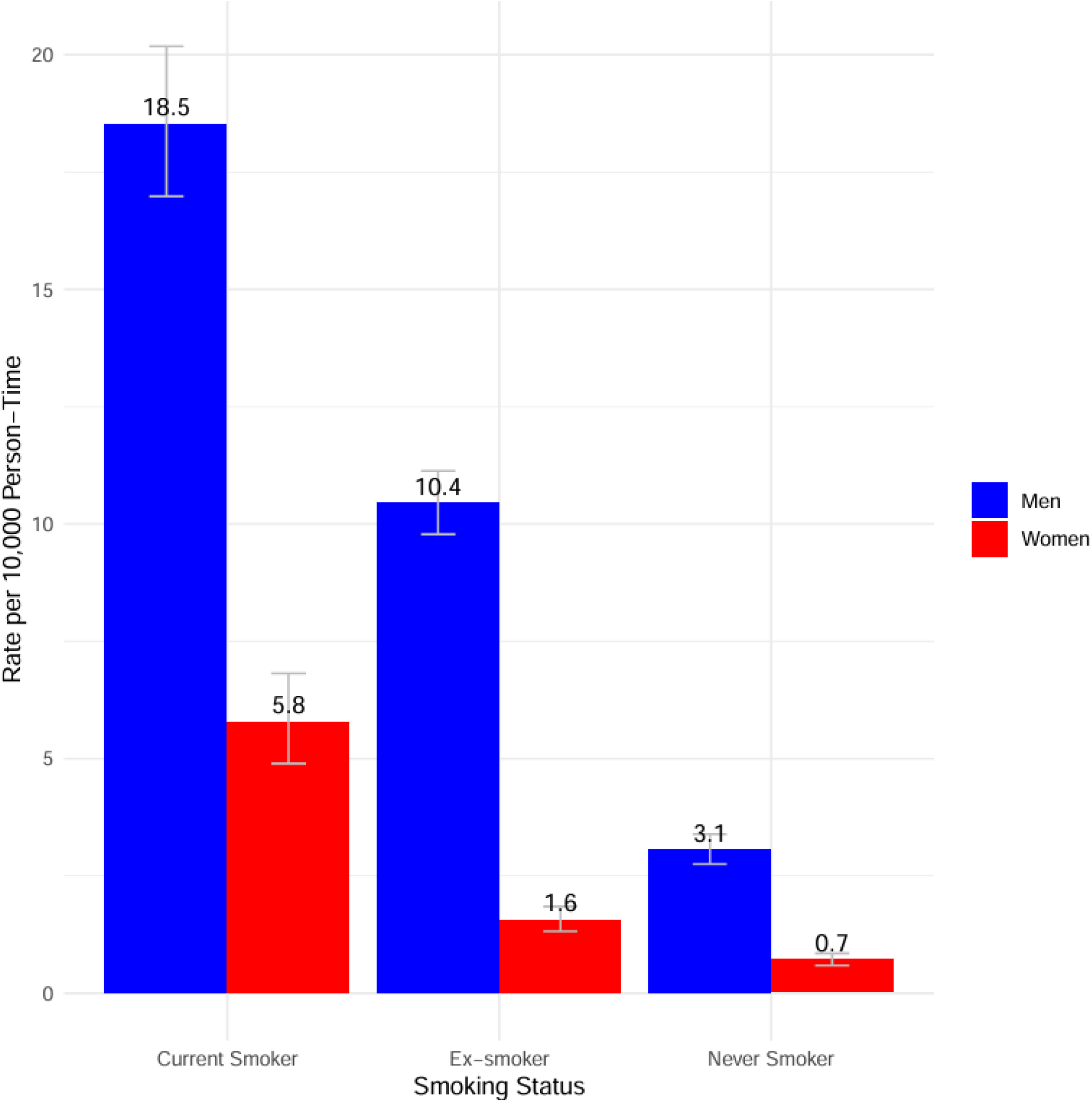
Rates of incidence of AAA in the UK Biobank cohort by smoking category for men and women.

The primary adjusted HR for current smokers versus never smokers in men and women for incident AAA are HR 3.78 and 6.60 respectively (Table 1). The test for sex interaction was highly significant (HR 0.57, 95%CI 0.45-0.72). The data used in these analyses are shown in Table 2.

The comparable HRs for incident ASCVD in men and women, for the same cohort, in current smokers versus never smokers were 1.82 [95%CI 1.74,1.90] and 2.34 [95%CI 2.19, 2.51] respectively. The test for sex interaction was highly significant but weaker than for AAA (HR 0.78, 95%CI 0.72-0.84).

### Investigation of potential modifiers of the sex-specific effects of smoking for incidence of AAA in the UK Biobank cohort

Neither of the sequentially adjusted comparisons of the risk of AAA in current smokers versus ex-smokers showed an important amelioration of the sex-specific difference in primary adjusted HRs, although the lower numbers in the second sequential analysis led to much wider confidence internals (Figure 3). The details of all these additional covariates are shown in Supplemental Table S1.

**Figure 3.**
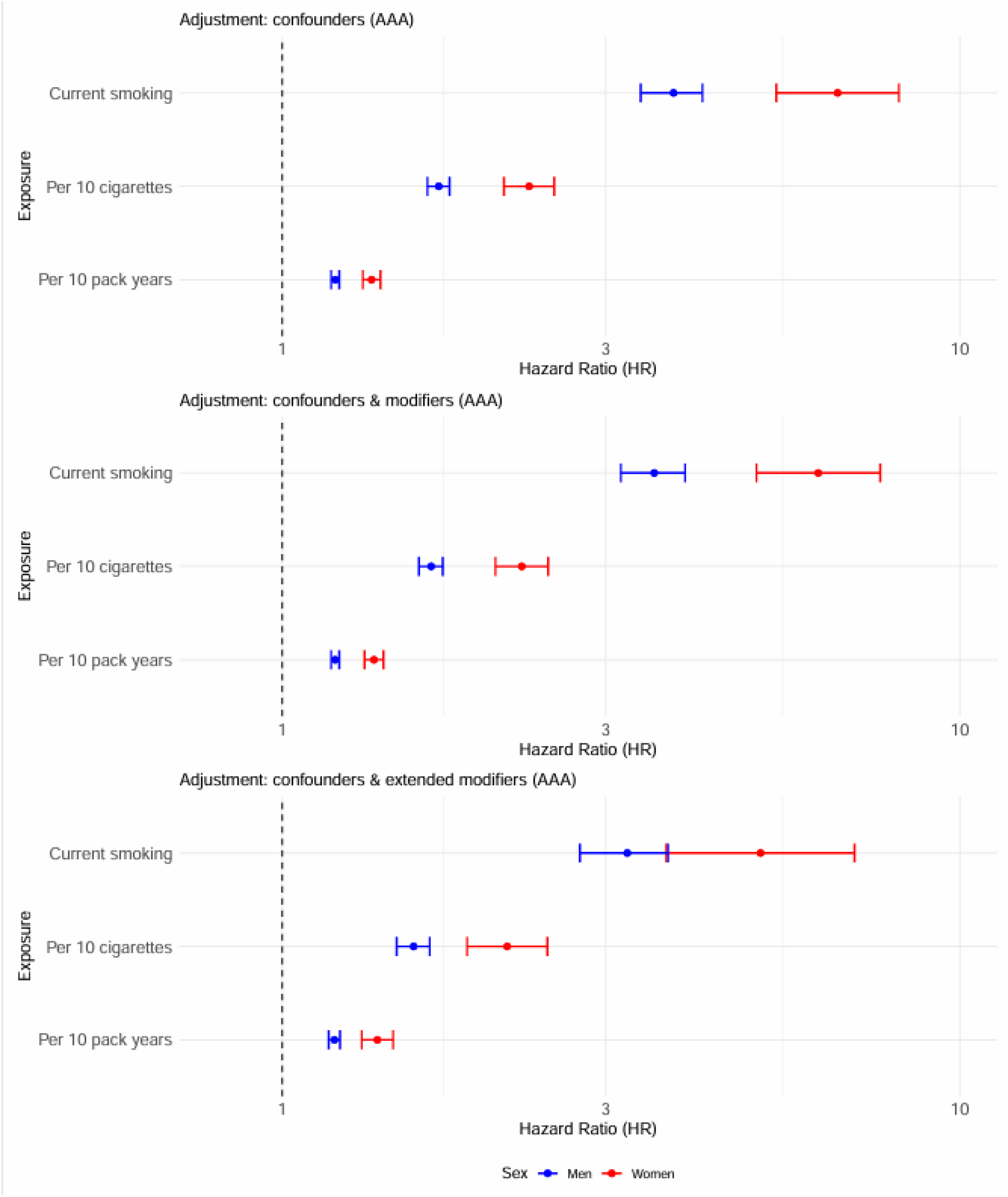
Forest plot showing the similar sex-specific differences in risk of AAA in current versus never smokers from UK Biobank for analysis by number of cigarettes smoked per day (in multiples of 10) and by pack-year history (multiples of 10).

Next, we considered the effects of the intensity and extent of smoking habits, The forest plot in Figure 3 shows the impact of number of cigarettes smoked per day and pack-year smoking history, with the number if cigarettes per day showing a stronger sex-specific difference than pack-year smoking history: test for sex interaction in the primary adjusted analyses HR 0.736 [95%CI 0.672, 0.865] and 0.884 [95%CI 0.858, 0.931] respectively. For comparison the impact of smoking habit on the sex-specific risks of ASCVD are shown in Figure 4: the effects of number of cigarettes per day also appeared stronger than pack-year history (Figure 4): test for sex interaction in the primary adjusted analyses 0.848 [95%CI 0.814, 0.884] and 0.922 [95%CI 0.909, 0.926] respectively. A comparison of splines that allow for non-linearity confirmed higher relative risks of current smoking for AAA in women compared to men (Supplemental figures S5 and S6). For both number of cigarettes/day and pack-years, the HR for AAA increased more steepply in women and increasing exposure to >30 pack-years or smoking >15 cigarettes/day had greater effect on increasing the risk of AAA in women.

**Figure 4.**
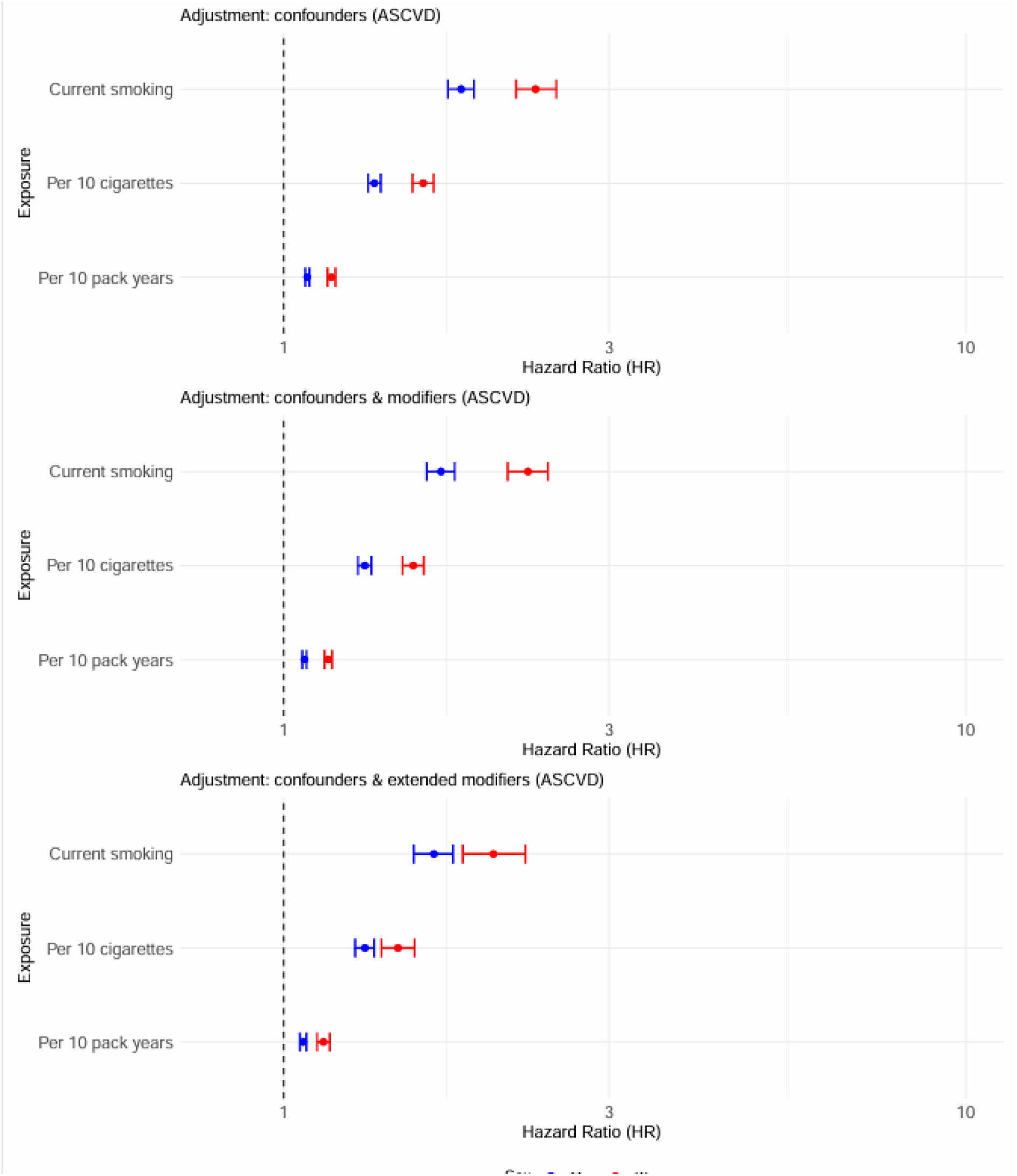
Forest plot showing the similar sex-specific differences in risk of ASCVD in current versus never smokers from UK Biobank for analysis by number of cigarettes smoked per day (in multiples of 10) and by pack-year history (multiples of 10).

To further investigate the role of inflammation, analyses were stratified by CRP concentrations below 2 mg/L and ≥2 mg/l indicated a weak potentiation by CRP. The HRs for the risk of incident AAA associated with current smoking at baseline are shown in Table The sex-specific difference remained almost unchanged in the lower CRP stratum.

## Discussion

The meta-analysis consolidates and extends the evidence from previous reports, putting improved precision on the relative risk of AAA in current smokers, which is almost twice as high in women versus men. The sex-specific relative risk ratio of smoking for AAA was much higher (1.78 fold) than the more modest 1.25-fold increase for ASCVD observed previously(10) or in the present UK Biobank cohort (1.29). This study is the first to explore potential modifiers that might help explain mechanisms underpinning the higher relative risk of smoking in women versus men. There was no evidence to support our hypothesis that this was mediated by inflammation or any other covariate investigated and the sex-interaction was still notable by smoking habit and history. The rate of AAA incidence was very low in female non-smokers (0.7 per 10,000 patient years) in the UK Biobank cohort. A very low incidence in non-smokers could contribute to the high relative risk associated with smoking in women, since AAA is rare in women who have never smoked – but the same was true in previous studies. Other possible contributory reasons for the high relative risk of AAA in female smokers include the much lower use of cardiovascular risk prevention medications in women or that men who have never smoked are at higher risk of AAA due to their lower levels of HDL-cholesterol and higher blood pressures.

The relative risk of current smoking for AAA in women varied from 0.82 to 2.69 in the different studies, pooled relative risk of 1.78 [95%CI 1.32 to 2.38], with high heterogeneity. Reasons for the high heterogeneity observed might be related to underlying population and ethnic differences, the varying study designs (cross-sectional or longitudinal), the mode of AAA ascertainment (screening in some studies or record-linkage clinical incidence in others) or even the definition of AAA (aortic diameter either 3.0 cm or 3.5 cm in in screening studies with various planes of measurement). In the UK Biobank cohort many of the AAA in men could have been detected through the national screening programme, where the majority of AAAs detected are small and in the early phases of development(19). Women are not offered screening and therefore incident AAAs were likely to have reached a more advanced stage of development. Since, the relative risk of smoking for AAA in women in the UK Biobank cohort was similar to that in other studies, based either only on screening or only on data linkage, the possibility of different stages of AAA development in men and women seems an unlikely explanation for the sex disparity observed.

Smoking also is known to have an increased risk for the early onset and severity of chronic obstructive pulmonary disease and ASCVD. Our first hypothesis for the very high excess risk of smoking for AAA in women was that it was mediated by inflammation. Indeed, baseline CRP concentrations were higher in women than men, but including CRP as a covariate in regression models or stratifying analyses by baseline CRP concentration had no significant on the relative risk of smoking in women versus men. Further, including alkaline phosphatase (another inflammatory marker) and white cell count did not attenuate the sex-specific effect of smoking on AAA incidence.

Further analysis of smoking in the UK Biobank cohort demonstrated a similar relative sex-specific disparity in risk of AAA by number of cigarettes/day (higher in women) but slightly less disparity for total smoking exposure (pack-years, which was lower in women). A similar trend was observed for ASCVD. This could suggest that smoking more cigarettes/day might contribute to the very high risk of current smoking for AAA in women. Previously, a small case-control study of smokers with AAA compared with smokers with occlusive aortic atherosclerosis identified depth of inhalation as a potential risk factor for AAA(20) and an increasing amount of daily smoking also has been associated with an increased risk of AAA in men(21). There is ample evidence of genetic risk factors for smoking as well as evidence of sex-differences in the neural processing of cigarette smoking(21). Therefore, a fruitful approach to understanding the mechanisms underlying the sex-specific effects of smoking in cardiovascular disease might be genetic investigation of the nicotine signalling pathways in the brain. This is an area where sex-specific differences in dopamine signalling pathways are increasingly important in neuropsychiatric disorders, including addiction, with sex-specific differences in dopamine receptor distribution, density and function(22).

Sex-specific endocrine interactions are another potential contributor to the sex-specific differences in the risk of smoking for cardiovascular disease. There is a dose-dependent effect of pack-year history of smoking on risk of early menopause, which is likely to accelerate biological aging in women(23) The mechanisms underlying this effect on early menopause are various but direct toxicity of smoking on ovarian follicles is supported by the dose-dependent effect of smoking, (number of cigarettes smoked,) to reduce Mullerian hormone concentrations(24).

Numerous biological pathways have been associated with AAA development spanning molecular damage to the aorta from the luminal thrombus to medial degeneration to neovascularisation and immune cell infiltration in the adventitia. Many of the specific cell and molecular pathways have been elucidated in animal models. Some of these models incorporate cigarette smoke exposure to mimic the human disease(25). The data presented here, reinforce the need to use both male and female animal models and analyse their data separately.

There are several limitations to our study. Only 6 studies contributed to the meta-analysis, too few for examining the risk of publication bias. Three studies, including UK Biobank, were longitudinal but only used baseline data for their analyses of the risk of smoking(3, 4, 18). None of these longitudinal studies made allowance for the competing risk of death from other causes. It is important to recognise that UK Biobank reports on a healthier than average population and in which there may be many unconsidered or unmeasured confounders(26). A further limitation of the UK Biobank analyses is that both smoking and other covariates data were only available at baseline.

In conclusion, the data here indicate that smoking is a much stronger relative risk factor for AAA in women than men, perhaps underpinned by the observation that AAA is very rare in women who have never smoked. This observation should impact future experimental studies as well as focusing on the need to both improve smoking cessation efforts in women and, particularly, to re-evaluate AAA screening for women smokers.

## Abbreviations used

AAA: Abdominal aortic aneurysm
ASCVD: atherosclerotic cardiovascular disease
CI: confidence interval
BP: blood pressure
CRP: C-reactive protein
FEV1: forced expiratory volume in 1 second
FVC: forced vital capacity
HR: hazard ratio
RR: relative risk.

## Acknowledgements

This study would not have been possible without the participants, investigators and funders.

